# For She had Eyes and Chose Preprints -– Social Media Impact, Citation and Publication rates of Ophthalmology preprints

**DOI:** 10.1101/2022.10.25.22281523

**Authors:** Rahma Menshawey, Esraa Menshawey, Maryam Massoud

**Affiliations:** Kasr al Ainy, Cairo University Hospitals, House Officer; Kasr al Ainy, Cairo University Medical School, Student

**Keywords:** Ophthalmology, preprints, COVID19, publication, medrxiv, biorxiv

## Abstract

**Background:** Preprinting, is the sharing of non-peer reviewed, unpublished scholarly manuscripts. Across many fields of medicine, an exponential rise of manuscripts being posted to preprint servers has been observed. This has exploded during the COVID19 pandemic where early dissemination of information was critical, or where COVID19 priority disrupted the publication dynamics and priorities in other areas. We examined the characteristics of ophthalmology related preprints in this study.

**Methods:** We searched the bioRxiv and medRxiv servers for preprints relating to field of ophthalmology. Preprints were screened by title and abstract to ensure they were related to the field of ophthalmology. Outcomes included number of tweets, upper bound followers of tweeters, number of citations, news outlets reports and dates posted/published. If a preprint was published the same outcomes were collected for the published version to allow for comparisons, as well as journal publisher and cite score.

**Results:** After screening, a total of 720 preprints met our inclusion criteria. 420 of the preprints went on to be published. The publication rate of the preprints was 58.3%. The median number of tweets received on preprints was 3.5, IQR 5.5. 98.75% of preprints were tweeted about. Citation rate was 40.14%. The average number of days from date posted on server to date published was 180.5±124 days. Regression analysis revealed that tweets as a preprint predicts tweets as a published article, P=0.0135. citations as a preprint predicts citations as a published article, P<0.001. The average cite score was 8.5±6.2 among those preprints that we published in journals. 16.66% of papers were published by Elsevier (n = 70). 3.19% of preprints were COVID19 related, with significant differences found between them and non-COVID19 preprints in terms of number of tweets and number of citations.

**Conclusions:** Ophthalmology preprints are increasing across preprint servers. This maybe to bypass publication times and allow early dissemination of work, as well as increase visibility and citations. We identified that preprint citations and tweets predict published version citation and tweets.

## Introduction

The COVID19 pandemic has greatly impacted scientific and medical publication dynamics. Some medical fields have faced reduced research funding and publication, and redistribution of resources away from non-COVID interests(Raynaud et al., 2021) The field of Ophthalmology uniquely experienced a rise in non-COVID19 publication during the year 2020 (Reitinger et al., 2021).

There remains a clear need for the quick dissemination of information during the pandemic in order to quickly share vital information that may impact or guide clinical care(Nabavi Nouri et al., 2021). The need to over-come publication process wait times was also a challenge. Preprint servers provide a place where manuscripts of original research can be posted online (typically in sync with sending the manuscript off to a journal for publication). This allows for the sharing of the scientific work with the public and other scientists or experts as the manuscript undergoes formal peer-review – preprints do not count as publications (although they are given a unique Digital Object Identifier and may be cited), and they are not formally peer-reviewed(Hodel et al., 2022). Preprint servers may inadvertently provide a place for manuscripts that cannot otherwise be published, and some concerns have been raised regarding if they are able to withstand the review process and research integrity(Gopalakrishna, 2021). One study observed that preprints on the COVID topic had a publication rate of 5.7% and this draws concern on the scientific validity of these works. (Añazco et al., 2021)

Overall, preprint servers have benefits including establishing the priority of a work, promoting collaboration and feedback on the work, and being open-access(Sarabipour et al., 2019). The purpose of this study is the determine the citation and publication rates, and social media impact of Ophthalmology preprints during the time of the pandemic.

## Methods

A search was conducted on two major preprint servers (BIORXIV and MEDRXIV) for Ophthalmology related preprints. MEDRXIV had its own Ophthalmology subject area containing all related preprints. Independent search on BIORXIV for the key word “Ophthalmology” was conducted. Temporal limits were set to Jan 1 2020 to Dec 31 2021 for the search on the Biorxiv server.

Search of the MEDRXIV Ophthalmology collection yielded 187 preprints, while BIORXIV yielded 1285 preprints.

Preprints were additionally screened by title and abstract or further reading to determine that the preprint can be classified as an Ophthalmology topic (done by EM, RM, and MM).

An Excel sheet was developed to collect data on the following outcomes:

NUMBER OF AUTHORS, COUNTRY OF ORIGIN, NUMBER OF TWEETS, COUNTRY WITH THE MOST TWEETS, UPPER BOUND FOLLOWERS OF TWEETERS, NEWS OUTLET POSTS, CITATIONS, TIMES ABSTRACT WAS READ, TIMES FULL PDF WAS DOWNLOADED, DATE POSTED, PUBLISHED OR NOT, AND COVID TOPIC OR NOT

*If* a preprint was also published, the following outcomes were also collected:

DATE POSTED, CITATIONS, TWEETS, UPPERBOUND FOLLOWERS OF TWEETERS, NEWS OUTLET POSTS, PUBLISHER OF THE JOURNAL, CITE SCORE OF THE JOURNAL.

Country of origin was determined as the country of origin of the corresponding author.

The selected outcomes metrics could be determined directly on the journal itself if it provided these metrics, or if they hosted a metric tool such as PlumMetrix, or Altmetrics. Uniformly, all metrics for the preprints were either hosted by the preprint server or Almetrics, while multiple methods were used to derive these results for the published papers as journals means to report metrics was variable. Data search was conducted October 1^st^2022, and data collection was completed by the authors October 8^th^ 2022. Results were screened for duplicates, and duplicates identified were removed.

## Statistical Analysis

Analysis was performed using MedCalc. Quantitative data was represented as mean and standard deviation or median and interquartile range when appropriate. Data was explored for normality using Shapiro-Wilk tests. Regression analysis was performed to determine the relationship between two variables. Spearman’s rho was used to determine rank correlation when distribution was not normal. For non-parametric data, Mann-Whitney U test was used to compare variables between the 2 groups. Statistical significance was set at <0.05. **All statistical analysis was performed using MedCalc for Windows version 19.1 ((MedCalc Software, Ostend, Belgium)**

## Results

A total of 1472 preprints were identified between the servers (BIORXIV= 1258, MEDRXIV = 187). After screening and removal of duplicates (n=16), a total of **720** (BIORXIV = 536, MEDRXIV = 184) preprints met our inclusion for further analysis. Table 1 contains all the preprint characteristics. Table 2 contains the published preprint characteristics.

**Table 1:** Characteristics of Ophthalmology Preprints

| **Preprints (n=720)** | **Lowest value** | **Highest value** | **Mean + 95% CI** | **Standard Deviation** | **Median + 95% CI** |
| --- | --- | --- | --- | --- | --- |
| **Number of authors** | 1 | 51 | 7.6236<br>(7.2564 to 7.9908) | 5.019 | 6.0000<br>(6.0000 to 7.0000) |
| **Number of Tweets** | 0 | 269 | 8.4000<br>(6.9072 to 9.8928) | 20.40 | 3.5000<br>(3.0000 to 4.0000) |
| **Upper Bound Followers of Tweets** | 0 | 1428682 | 97462.4<br>(90596.8 to 104328.03) | 93113.2 | 100972<br>(97873.4 to 104198.08) |
| **PDF downloads** | 5 | 17491 | 529.36<br>(468.37 to 590.34) | 827.64 | 382.5<br>(362 to 403.6) |
| **Abstract Views** | 52 | 22302 | 1490.2<br>(1370.77 to 1609.73) | 1621.6 | 1068.5<br>(1019.8 to 1123) |

**Table 2:** Characteristics of Published Preprints

| **Published (n=420)** | **Lowest value** | **Highest value** | **Mean + 95% CI** | **Standard Deviation** | **Median + 95% CI** |
| --- | --- | --- | --- | --- | --- |
| **Number of Tweets** | 0 | 667 | 12.1689<br>(8.2935 to 16.04) | 37.7539 | 4<br>4 to 5 |
| **Upper Bound Followers of Tweets** | 0 | 1931498 | 43974.72<br>30257.23 to 57692.21 | 132346.08 | 7161.5<br>5334.04 to 9307.57 |
| **Citations** | 0 | 212 | 7.3<br>(5.74 to 8.88) | 16.29 | 3<br>(3 to 4) |

Preprints originated from 38 distinct countries. The countries that had posted the most preprints were the United States (335), China (63), United Kingdom (57) and Germany (56) (See **figure 1**). The number of preprints posted in each year are depicted in **figure 2** – no significance was found in this rank (Spearman’s Rho = 0.50, p=0.6667).

**Figure 1:**
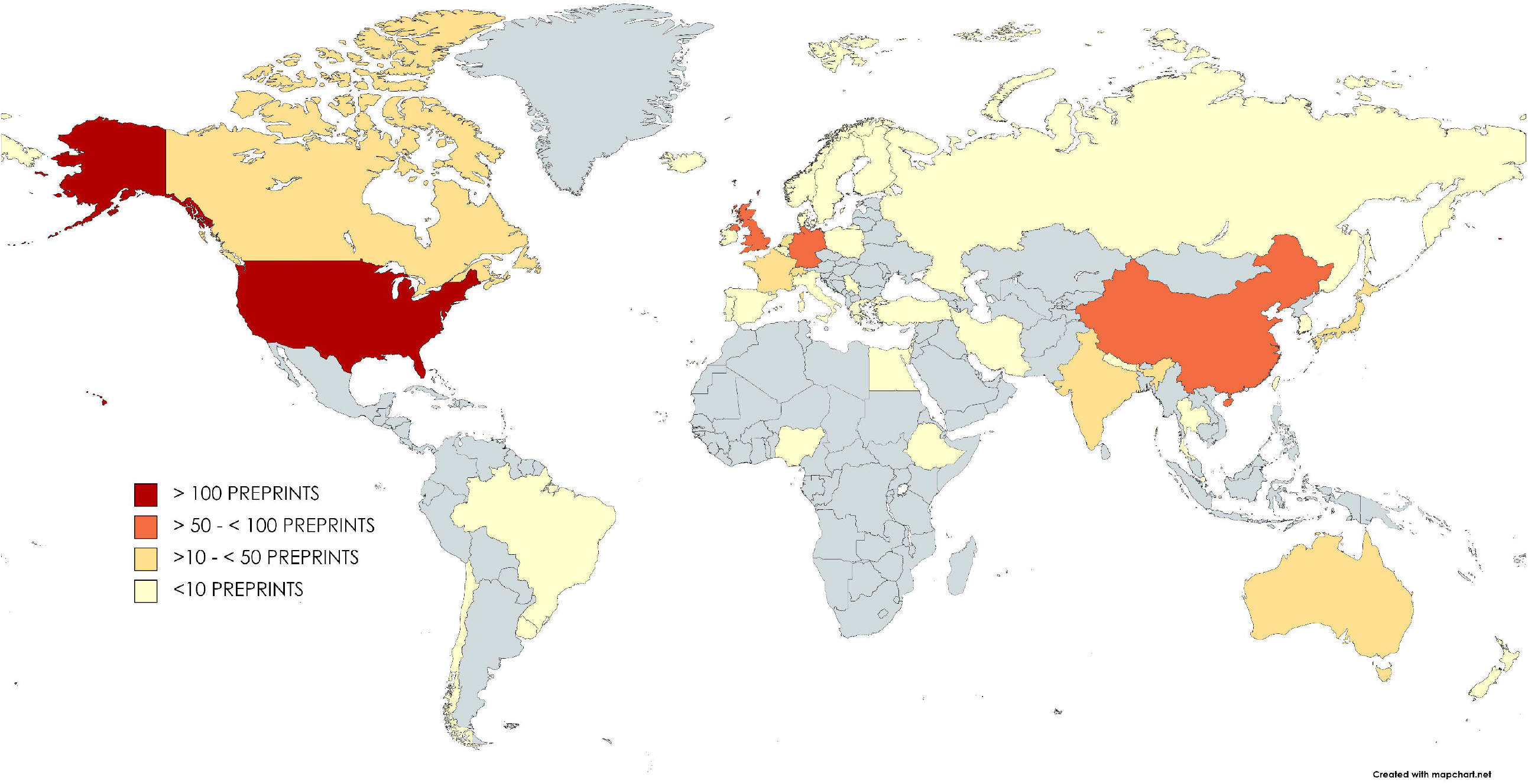
Map of Country of Origin of Preprints.

**Figure 2:**
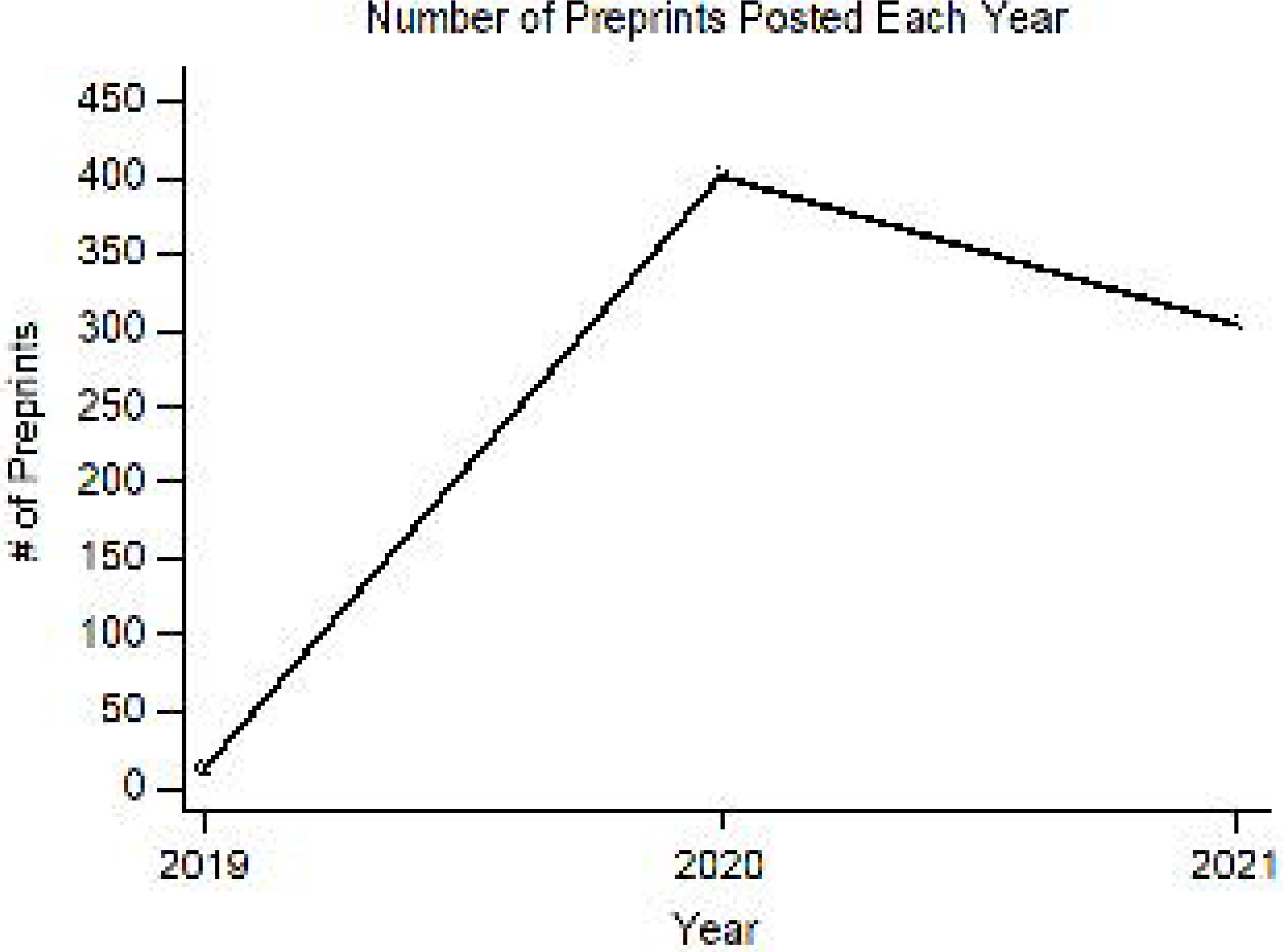
Number of preprints posted each year.

Of the posted preprints, 421 have been published so far; the publication rate for Ophthalmology preprints was 58.3%.

The median number of authors on preprints were 6, IQR 6. The median number of tweets received on preprints was 3.5, IQR 5.5. 711 of the preprints were tweeted about, or 98.75%. Countries that had the tweeted preprints are depicted in **figure 3**. The countries that had done the most tweeting were: the United States (491 tweets), United Kingdom (57 tweets), Singapore (20), Germany (12), and Australia (10). The number of tweets increased significantly over the years (Spearman’s rho = 0.250, P<0.0001)

**Figure 3:**
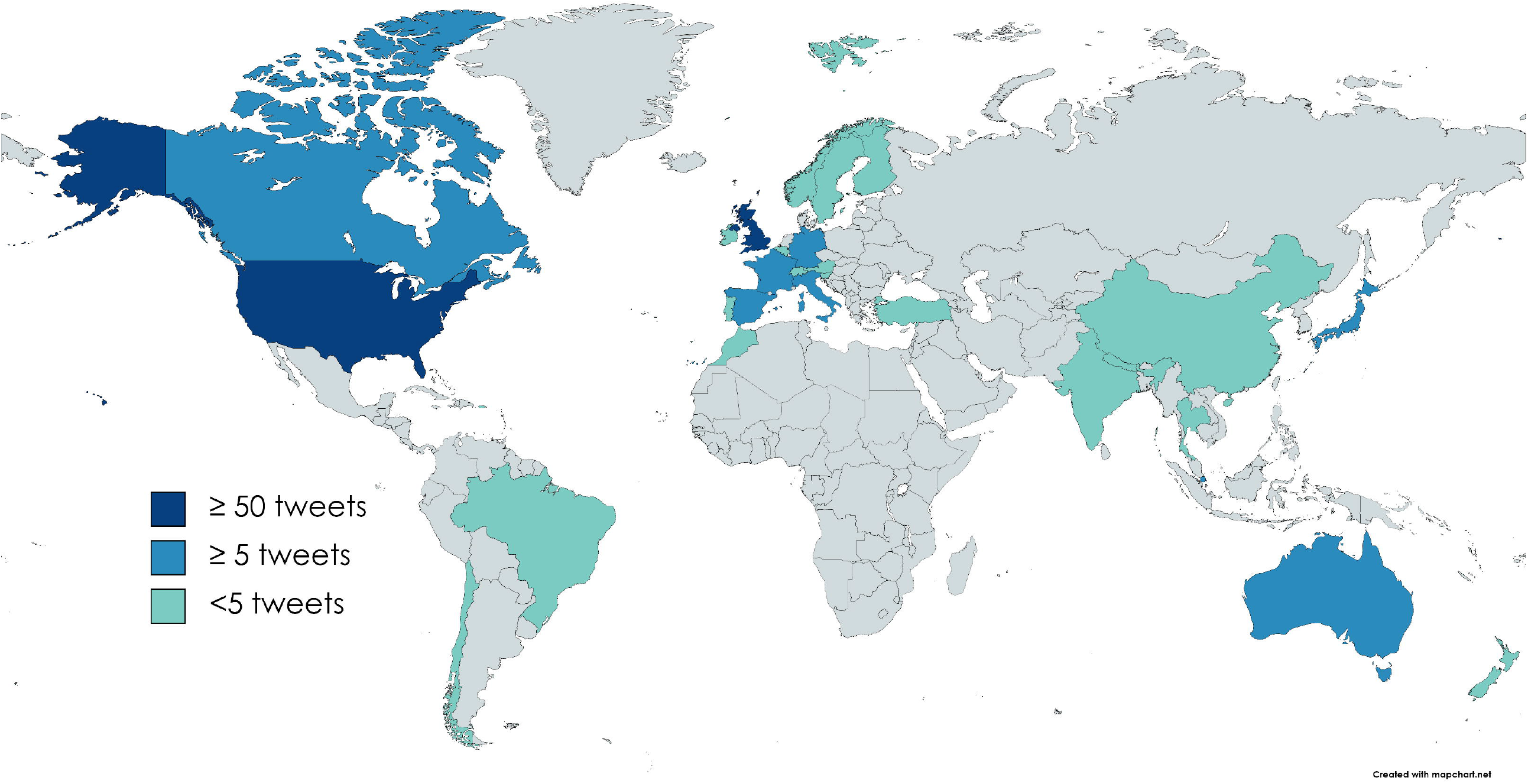
Countries that posted Tweets about the examined preprints.

The upper bound followers of tweeters (which may represent the potential number of people who have read the tweet(s)) median for preprint papers was 100972, IQR 100334.5. **Figure 4** depicts the type of the tweeters – the majority were members of public (92%).

**Figure 4:**
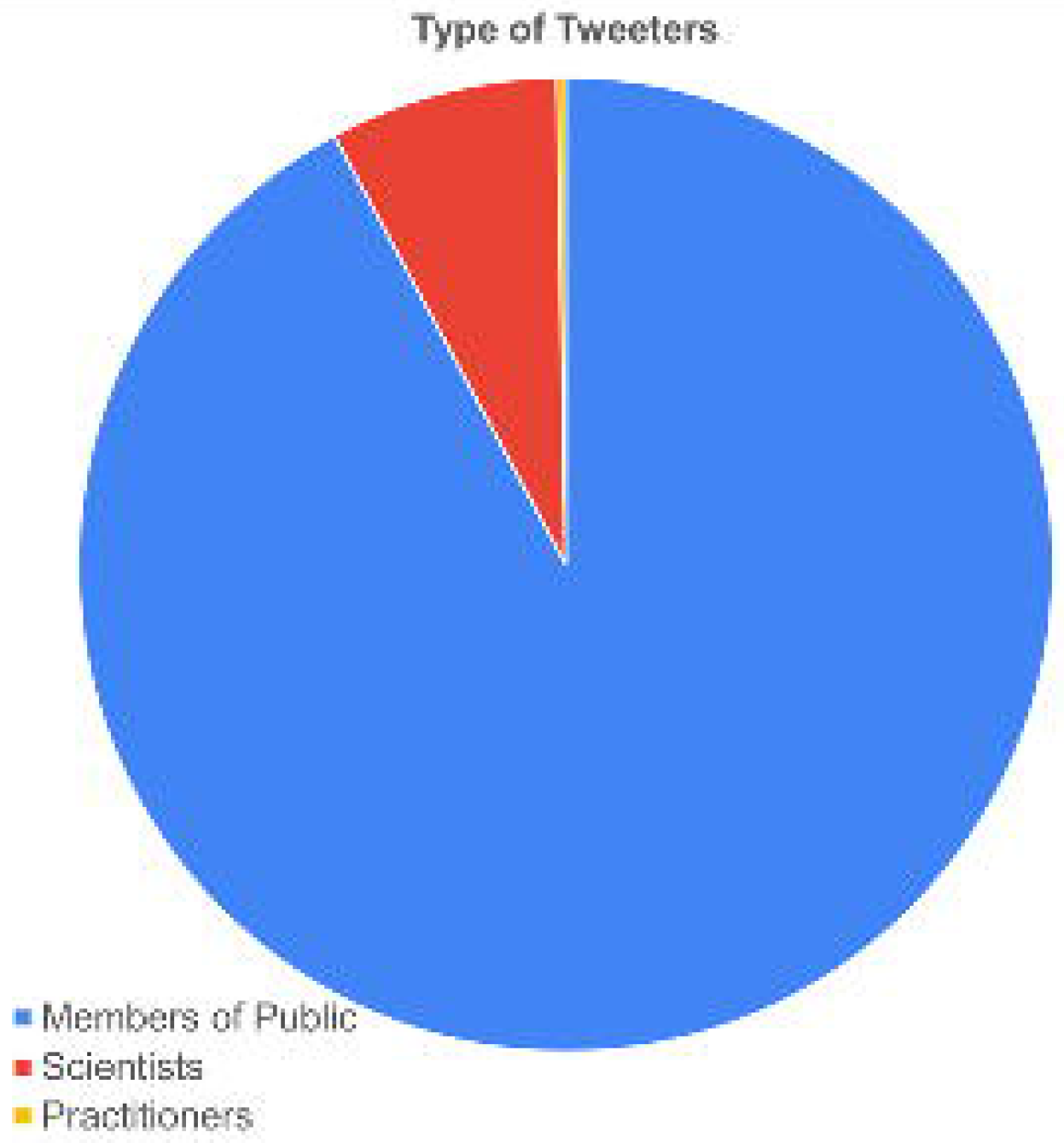
Type of Tweeters as determined by Altmetric.

Only 19 of all the Ophthalmology preprints were reported on in news outlets as determined by Altmetric (2.64%), with the lowest number of times being 1 and the highest number of times reported in the news being 88. The median number of news reports among them was 2, IQR 7. 63.16% of preprints reported on in the news originated from core Anglosphere speaking countries (United States, United Kingdom, Canada, Australia)

The median number of PDF downloads of the preprints was 382.5, IQR 313. The median number of abstract views of the preprints was 1068.5, IQR 842.

40.14% of preprints were cited (n= 289). The lowest number of citations was 0 and the highest was 1957. Among those that were cited, the median number of citations was 2, IQR 2.

Table 2 depicts the characteristics of preprints that went on to be published. A total of 421 preprints were identified as “published” (the preprint servers automatically connect/link to the published version). However, 1 article had to be excluded as even though it was listed as “published” by the server and on BMJ open (where it is allegedly published), the link was not connecting to the published work. External search by title on Pubmed, Google, as well as on BMJ open did not yield the paper in question, so for further analysis it was not included. The final analysis on 420 published preprints. The publication rate therefore of the Ophthalmology preprints was 58.33%.

The average number of days from date posted on server to date published was 180.5±124 days, or 5.91±4.07 months. The longest it has taken so far for a preprint to be published was 690 days, or 1.88 years. We observed 4 preprints that were posted to the servers after they were published in a journal, and this is against the purpose of the preprint servers and serves as a redundancy in the literature-1 additional preprint was posted 2 days before it was made publicly online.

The average number of citations of the published work was 7.3±16.29. The citation rate of the preprints overall was 39.3%. **Figure 5** depicts scatter diagram with regression line of regression analysis for published versus preprint citations. This result suggests that citations as a preprint predicts citations as a published article, P<0.001. The published papers were tweeted on average 12.16±37.75 times, with an average upper bound followers of users of 43974.72±134346.08 (this represents the possible number of people exposed to tweets about the published work). Regression analysis revealed that tweets as a preprint predicts tweets as a published article, P=0.0135 (see **figure 6**). Statistically significant difference was found between preprints citations and citations of the same preprints that went on to be published, P <0.0001 (See figure). No statistically significant differences were found between tweets of preprints versus published version of papers. P =0.00848.

**Figure 5:**
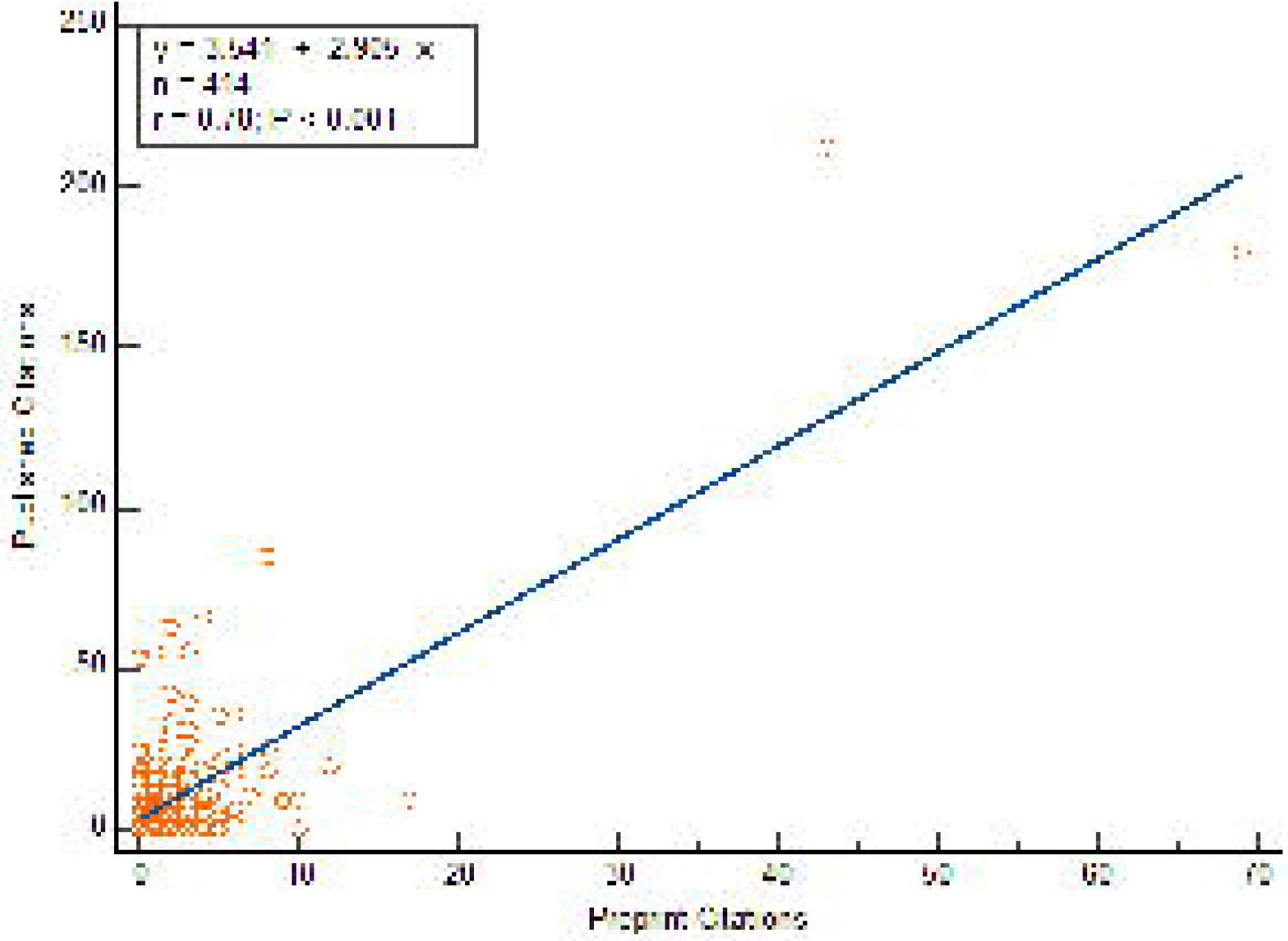
Scatter diagram with regression line depicting published version versus preprint number of citations.

**Figure 6:**
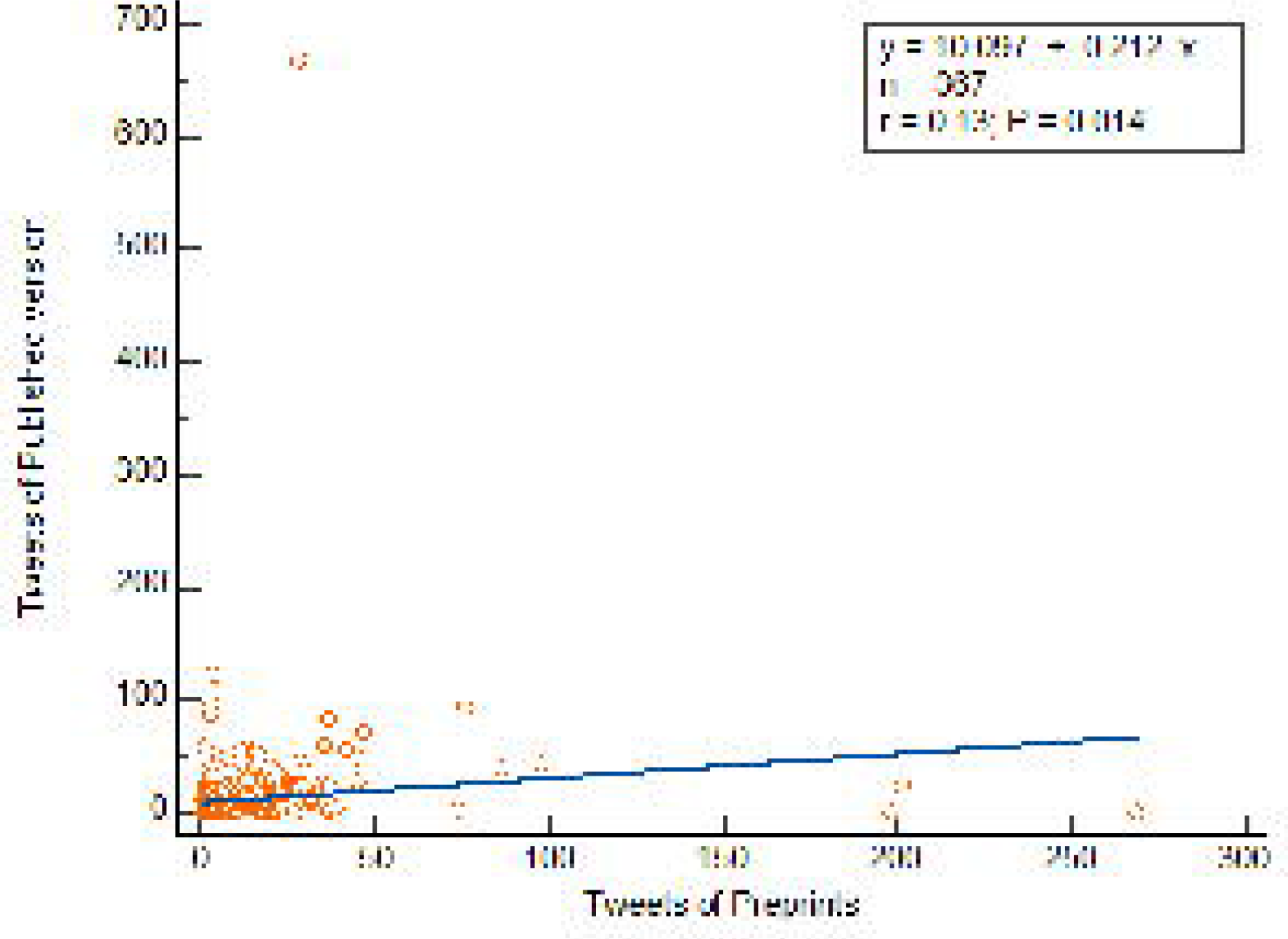
Scatter diagram with regression line for number of Tweets of preprint versus published version.

16.43% of published preprints were reported on in news outlets (n=69). The lowest number of news reports was 1, and the highest was 120. Cite score was an outcome we considered for the journals of the published works. Cite score represents the average number of citations of published articles in a journal. The lowest cite sore we observed was 1.3 and the highest was 70.2. The average cite score was 8.5±6.2 among those preprints that we published in journals. Publishers that had published work previously posted as a preprint are depicted in **figure 7**. 16.66% of papers were published by Elsevier (n = 70).

**Figure 7:**
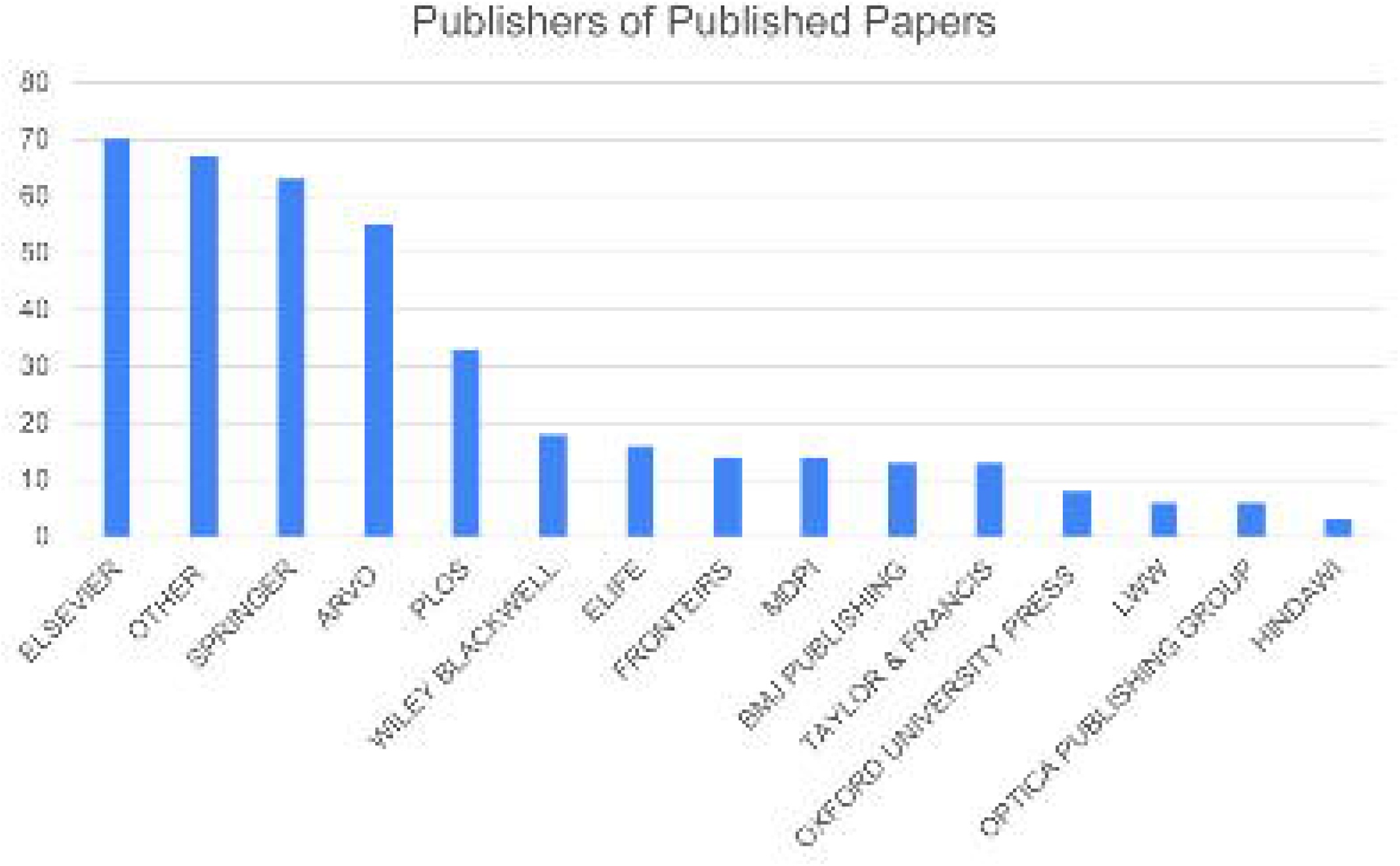
Publishers of journals that published preprint articles.

Finally, we identified 3.19% of Ophthalmology preprints were COVID19 related (n=23). The median number of authors was 6, IQR 5. Preprints originates from China (n=7), UK (n=4), USA (n=4) and others. 69.57% of COVID19 preprints were published (n=16). Median number of tweets was 8, IQR 39, all of which were tweeted by members of public. Median number of PDF full text downloads was 837, IQR 1475. Median number of citations was 4, IQR 10. Among the published preprints, the days between date posted and date published was a median of 119 days (or 3.91 months), IQR 373. Median number of citations of published articles was 8, IQR 19.5. Significant differences exist between the number of citations of COVID19 preprints versus non-COVID preprints (p<0.0001), see **figure 8**. Significant differences were also found between the number of tweets of COVID19 ophthalmology preprints versus non-COVID related preprints (p=0.0001) **see figure 9**.

**Figure 8:**
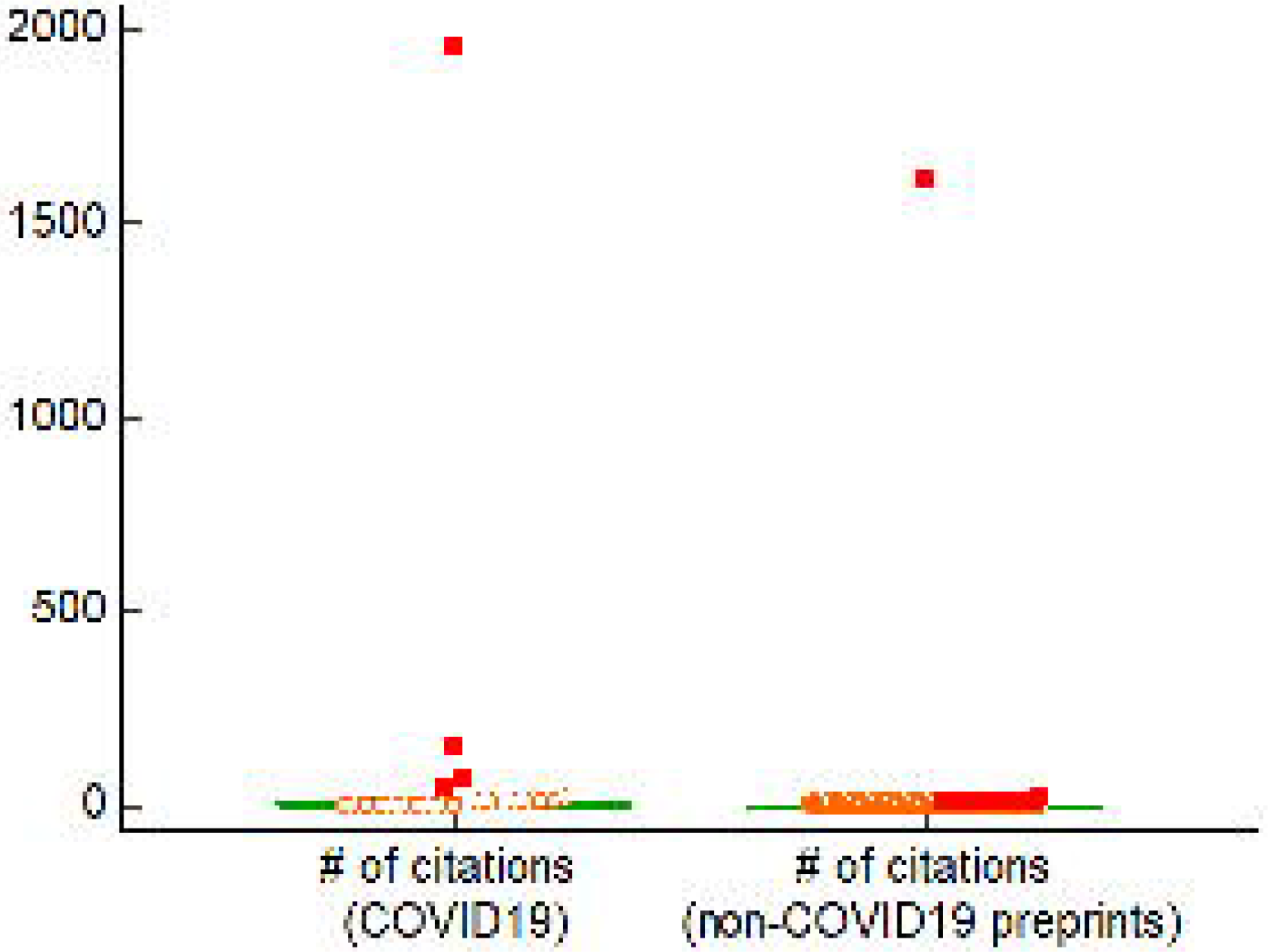
Significant differences found between # of citations of COVID19 preprints versus non-COVID19 preprints.

**Figure 9:**
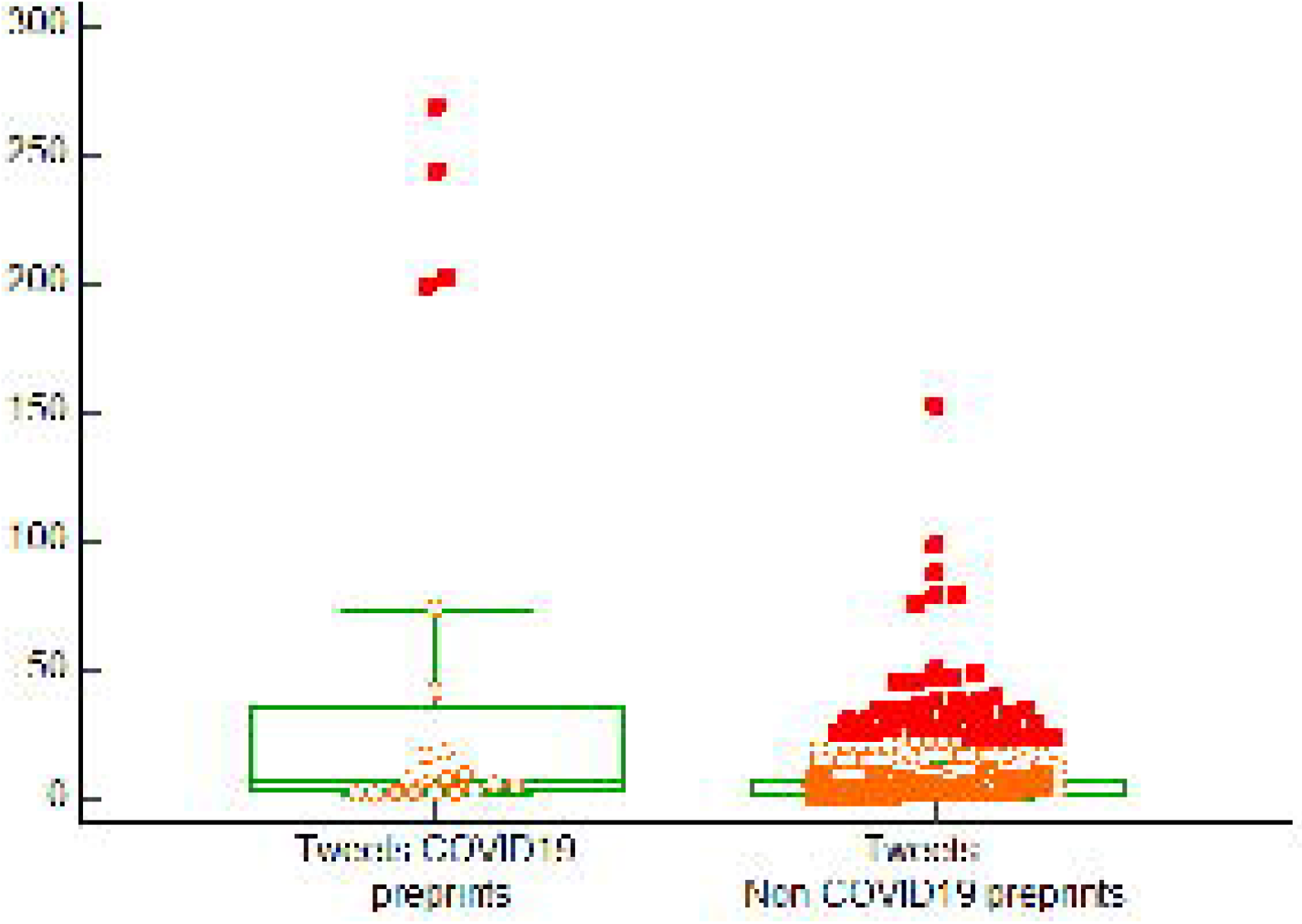
Significant differences found between Tweets of COVID19 preprints versus non-COVID19 preprints.

## Discussion

In this study we examined the characteristics of Ophthalmology preprints. We found that preprint servers provide a landscape whereby ophthalmology research can be posted in advance to publication in order to facilitate dissemination of information through casual reading (abstract view, pdf downloads), citations, social media spread (Twitter), and news outlets.

A preprint is a non-peer reviewed scientific manuscript that is uploaded to a platform, made available to the public(Kirkham et al., 2020). Posting one’s work has some benefits including early exposure, the ability to “claim” a research topic, and early constructive feedback. Preprint platforms provide a Digital Object identifier (DOI) and this allows the preprint to be cited by others(Kirkham et al., 2020). One major criticism of preprint servers is they allow work to be hosted on them which has not received formal peer review, some of which has not yet been published and calls to question the validity of the preprints hosted on them(Kirkham et al., 2020)(Malicki et al., 2020)(Añazco et al., 2021).

Studies on the forecasted publication rate of preprints in other areas (COVID19) revealed that 70% of preprints were published within 1 year(Gordon et al., 2022). Other studies found that two-thirds of preprints were eventually published(Abdill & Blekhman, 2019). In our study, we found that Ophthalmology preprints were published at a rate of 58.33%, and were cited at a rate of 39.3%. When determining the difference in time published date and preprint posted date, we discovered that average time between them was 180.5±124 days, or 5.91±4.07 months, for preprints that were posted across the years 2019 2020 and 2021. The longest time recorded for a preprint to be published was 690 days or 1.88 years. These findings establish the continued need for the existence of preprints servers as otherwise research which holds vital information that may be impactful to patient care is effectively backlogged in the publication process. Preprint servers provide a means for the immediate dissemination of research until such time that is formally peer reviewed and published.

The COVID19 pandemic caused drastic changes to the publication process to promote the dissemination of important information during this public health emergency. Preprints played a vital role in bypassing the publication times (although many journals aimed to provide prioritised peer-review and faster publication of COVID research). A study by Raynaud et al discovered that the COVID19 pandemic was associated with an 18% decrease in non-COVID related publications, as some editorial strategies included decreasing non-COVID publications and integrating COVID ones(Raynaud et al., 2021). These findings may further exacerbate publication times of non-COVID work as COVID research currently hold priority. This further supports the need for preprint servers as a viable platform to post one’s work until the time of publication or until some stabilization occurs in publication priorities. The COVID19 pandemic had directly influenced the dynamics of ophthalmology publications with one study identifying increased publications on oculoplasty due concurrent mucormycosis, as well as increased research output coinciding with COVID19 waves of infection in the Indian Journal of Ophthalmology(Gurnani & Kaur, 2022). Maximum citations were identified in general ophthalmology due to practise patterns and COVID19 influence(Kaur & Gurnani, 2021). Overall, there is a wide variation in time to publication among Ophthalmology journals as a whole; Dhoot et al identified a reduced number of days to electronic publication with higher impact factor journals(Dhoot et al., 2022). Even among COVID19 research, ophthalmology publications accounted for less than 0.5 percent of total articles (Forouhari et al., 2022). In our study, we identified significant differences between COVID19 ophthalmology preprints in terms of number of citations and tweets, versus non-COVID19 preprints. This resonates with the observations that COVID19 research was the priority.

Though we are the first study to examine ophthalmology preprints, other studies have analysed the characteristics of preprints in other specialties. One study examined pharmaceutical industry authored preprints and their social media impact of 498 preprints across several preprint servers. Median number of citations was 0, IQR = 1, while the median Altmetric score was 4(Subramanian et al., 2021). Overall, pharmaceutical industry preprints were increasingly being posted on preprint servers, cited as preprints and discussed on social media.

In trauma and orthopedic surgery, characteristics of preprints were examined by Hodel et al across 5 major preprint servers. They discovered that the number of preprints increased throughout the years, with a publication rate of 38.6%, and a mean time to publication of 8.7 months(Hodel et al., 2022). Likewise, we also identified an increase in the number of preprints throughout the years.

In the field of lipidology, an exponential rise of preprints was found across medRxiv and BioRxiv servers, while a general positive impact is perceived(Perera et al., 2022). In kidney disease research, it was observed by Vlasschaert et al that preprints in nephrology were also growing at an exponential rate nearly a 1 year delay in publishing clinical results(Vlasschaert et al., 2021).

Controversy continues to surround the posting of medical research on preprint servers prior to peer review with many calls to increase standards to prevent bad science that bypasses the peer review(Sheldon, 2018). Nonetheless, there is some appeal to posting work as a preprint first including early dissemination of research until the time that is published which may be prolonged if the research is not a priority especially in the setting of global health pandemic as the COVID19 pandemic. Preprints allow authors to establish the priority of their work onto a public onto a public record, as well as increase visibility to the work. Preprints in their own way provide a network type of “peer-review” as they can receive feedback publicly from a wider audience(Fry et al., 2019). Additionally, the main criticism against preprints, the lack of peer review, does not itself reconcile the fact that peer-review is imperfect with many examples of errors and misconduct identified years after publication in a journal. Looking forward, preprint servers could consider a threshold of time for which they host preprints in order to address the valid concern that unpublished work could represent research that does not withstand scientific scrutiny. This could be achieved by further analysis of dates posted versus dates published to place an upper limit on how long a preprint remains on a server. However, we noticed several instances were published work was then reposted to the preprint server and this represents a redundancy in the literature. Additionally, some dates had very few days in between that could not be excused as peer-review time. Therefore, we observed that preprint servers are being misused in this regard. Furthermore, Gehanno et al identified that preprints were being cited when the published version exists and this is faulty practise as the published version often has faced changes and revisions compared with the preprint version(Gehanno et al., 2022). We did observe significant differences in both the number of citations and Tweets of COVID19 related preprints versus non-COVID19 preprints – this suggests the priority and attention on COVID19 related work in both social media attention and citations compared with all other types of Ophthalmology research.

There are limitations to our study. Firstly, we utilised only two preprint servers, although they are major preprints servers. Future studies should examine other preprint servers. Time is an inevitable confounding bias in this study. Some studies that examine the characteristics of preprints have done so on a short timeline that does not accommodate potential delays in publication times. The studies we examined were posted at most in the year 2021, giving substantial time to determine a more accurate picture of the publication rate of preprints. This study is the first to examine the characteristics of Ophthalmology preprints on 2 major servers and describe their impact in terms of citations, news reports, and social media impact. This study provides a baseline data for Ophthalmology preprints providing a view into the landscape of work posted onto the preprint servers and the utility of preprints in Ophthalmology research dissemination dynamics.

## Conclusion

As publication dynamics and priorities continue to evolve, this work serves to highlight the importance and utility of ophthalmic preprints in continuing the dissemination research when those priorities do not majorly involve the ophthalmic field. Our findings suggest that preprint servers provide the means for early citation, and attention to research, and serve as a way to have an indisputable claim over one’s scientific discoveries. More and more ophthalmology research is being posted onto preprint servers and this could be reflective of the need to bypass prolonged publication delays, or increase readership and citation of research.

## Data Availability

All data produced in the present study are available upon reasonable request to the authors

## Ethics Declarations

### Ethical Approval and Consent

None required

### Consent for publication

Not Applicable

### Availability of data and material

Available upon reasonable request to the corresponding author

### Authors contributions

RM devised the research idea and design. RM EM MM participated fairly in data collection, writing the manuscript and final revision. RM conducted the statistical analysis on this study.

## Acknowledgements

None

